# Innovations, recipients, and contexts in the implementation of pediatric neurocritical care programs: a multisite qualitative study refining i-PARIHS

**DOI:** 10.64898/2026.10.13.26365431

**Authors:** Mia T. Vogel, Jessica S. Wallisch, Bradley J. De Souza, Pradip P. Chaudhari, Asmaa Hassan, Anvi Jampani, Katherine M. Steffen, Jose A. Pineda

## Abstract

**Background:** Pediatric neurocritical care (PNCC) has emerged as a distinct field, and many hospitals have moved to establishing dedicated PNCC programs. How practice change is implemented within these programs is not well understood. This study had two objectives. First, we sought to understand the barriers and facilitators of practice change in PNCC from the perspectives of the multiprofessional clinicians who deliver it. Second, we sought to characterize how well the integrated Promoting Action on Research Implementation in Health Services (i-PARIHS) implementation determinant framework captures them.

**Methods:** We conducted semi-structured interviews with a purposive sample of clinicians involved in PNCC at three U.S. academic medical centers. The interview guide was developed using i-PARIHS to organize barriers and facilitators of practice change across the innovation, recipients, context, and facilitation constructs. We applied framework analysis, combining deductive i-PARIHS, study-specific, and inductively derived categories. Two coders independently indexed the transcripts and reconciled them to consensus, with first-author adjudication. Preliminary site-level findings were validated with participants.

**Results:** We interviewed 33 clinicians: 24 physicians, 7 nurses, and 2 advanced practice providers. Most determinants of practice change mapped to existing i-PARIHS constructs, but three were not adequately captured by the framework as currently specified. The most consequential was an interprofessional authority gradient that determined whose preferences prevailed when intensive care, neurology, and neurosurgery disagreed. The other two were clinician autonomy, which served as the lens through which the acceptability of a change was judged, and the affective state clinicians bring to practice change. Receptivity depended more on recipients’ values, prior experience, and capacity to absorb change into a demanding workflow. Facilitation was variable, frequently under-resourced, and dependent on credibility across professional groups. We translated these findings into three propositions linked to implementation strategies and outcomes.

**Conclusions:** Participants attributed practice change in PNCC more to the recipients of change, the multiprofessional context, and the availability of facilitation than to the strength of evidence for a given practice. i-PARIHS captured most barriers and facilitators well but may be adapted in this setting. Credible, resourced, authority-sensitive facilitation may improve the prospects for sustained practice change as institutions develop PNCC programs.

**Contributions to the literature:**

- Determinant frameworks are widely used but rarely revised once published; this study supports three i-PARIHS framework modifications potentially applicable to the general pediatric ICU and PNCC programs regarding clinician autonomy, the affective state clinicians bring to change, and the interprofessional authority gradient.
- We identify authority-sensitive facilitation as an important driver of sustained practice change amongst multispecialty teams.
- We offer three i-PARIHS elaborations with linked implementation outcomes and implications for selecting implementation strategies for teams with shared authority across professions.

## Background

Over the past three decades, Pediatric Neurocritical Care (PNCC) has developed into a distinct subspecialty field (1–3). Acute neurologic illness and injury are common in the Pediatric Intensive Care Unit (PICU), and they profoundly impact how children live afterward (4). In a U.S. analysis, at least one critical care intervention was recorded in 23% of children hospitalized with a primary neurologic diagnosis (5). The conditions involved (traumatic brain injury, stroke, status epilepticus, central nervous system infection, hypoxic-ischemic brain injury, and postoperative neurosurgical conditions) carry substantial risks of death and long-term neurodevelopmental morbidity (2, 5, 6). As mortality from pediatric critical illness has fallen, neurodevelopmental outcomes of PICU survivors have increasingly become a focus of critical care efforts (3, 6).

Drawing on the adult neurocritical care experience, in which dedicated units and specialized providers have been associated with better outcomes (3), pediatric institutions have established neurocritical care programs to coordinate multidisciplinary care that include deployment of evidence-based pathways and standardization of neuromonitoring and neuroprotection protocols (1, 7–9). These efforts have been associated with improved outcomes after severe traumatic brain injury and in other neurocritical conditions (10–12). Building a PNCC program requires implementing new and often complex practices within an existing, demanding intensive care environment, and the processes by which such change is achieved and sustained are complex and not completely understood (13).

Implementation science exists to help decipher those processes, and its theoretical approaches are widely sorted into process models, determinant and evaluation frameworks, classic theories from other fields, and implementation theories developed from the ground up (14–17). Among determinant frameworks, the integrated Promoting Action on Research Implementation in Health Services (i-PARIHS) framework holds that successful implementation is a function of the innovation, its recipients, and the context in which it is introduced, with deliberate facilitation of the process that integrates all three (18–22). i-PARIHS, and its predecessor PARIHS, have been applied across many health settings (18, 21, 23), and the developers have explicitly invited empirical work that tests and refines it (18, 24). The closest precedent for use of i-PARIHS in the PICU characterized blood product transfusion practice change across eight U.S. PICUs and concluded that the i-PARIHS framework was useful for identifying implementation determinants but would benefit from further elaboration of its facilitation subconstructs (22). i-PARIHS was therefore a suitable framework for use in PNCC, a similar setting, building on that previous work (25).

In this study, we sought to (1) understand the barriers and facilitators of practice change in PNCC from the perspectives of multiprofessional clinicians who deliver it and (2) characterize how well i-PARIHS captures practice determinants in the PNCC environment. We report a multi-site qualitative interview study and identify both the determinants of practice change that i-PARIHS organizes well and those for which elaboration would improve its fit.

## Methods

### Study design and reporting

This was a qualitative, interview-based investigation across academic medical centers designed to characterize determinants of practice change and to inform the development of PNCC programs at the participating sites. It is reported here in accordance with the Consolidated Criteria for Reporting Qualitative Research (26); the completed checklist is provided as Additional file 1. The study was approved by the Institutional Review Board at University of California, Los Angeles (Study # 23-000248). Verbal informed consent was obtained from all participants prior to their interviews.

### Object of implementation

The object of implementation in this study was evidence-informed PNCC practices or guidelines, and the organizational arrangements through which academic children’s hospitals deliver coordinated neurocritical care, for example neuromonitoring and neuroprotective clinical practice pathways, and the team structures that support them. Analysis examined determinants that recurred across practice changes rather than a specific practice. The guide is provided as Additional file 2. Where a determinant functioned differently for a discrete practice than for program-level organization, we note the distinction.

### Setting and participants

Interviews were conducted with providers at three U.S. academic hospitals, referred to here as Site A, Site B, and Site C, selected to represent institutions at differing stages of organizing their PNCC efforts. Two were stand-alone children’s hospitals; all were interested in developing a more formalized PNCC program within their PICUs. We used a purposive sampling strategy to obtain perspectives across the full range of professional roles involved in PNCC (27). A site representative assisted with identifying and recruiting eligible participants. Sampling continued until the team judged that further interviews were unlikely to alter the determinant structure (28).

Interviews were conducted by a member of the study team with qualitative research experience [M.T.V.] whose disciplinary background is in public health, social work, and implementation science. Interviews were conducted using a video-conferencing platform, audio-recorded, transcribed verbatim, and de-identified.

### Interview guide

The semi-structured interview guide was developed from the i-PARIHS framework as applied by Steffen et al. (22). It addressed the working environment for change in the ICU, recent PNCC practice changes, receptivity to change, decision-making, the use of guidelines, and perspectives on the development of a PNCC program. Initial questions asked participants to describe their role within the institution and PNCC activities, and a recent practice change as concrete entry points. We then explored barriers, facilitators, and the process of change.

### Data collection period and participant validation

Interviews lasted approximately 25 to 85 minutes (site medians 53 to 59 minutes). Interviews were conducted from 2023 to 2024, and site-level findings were shared with participants in October 2025 for validation and to confirm that the findings remained an accurate representation of the care environment (26, 29).

### Analysis

Framework analysis was applied to test and extend the i-PARIHS framework. This approach allowed us to chart and synthesize transcript-level data against an explicit analytic structure, keeping this report methodologically parallel to the prior PICU i-PARIHS study it builds on (22, 30, 31). The approach is deductive but allows for inductively derived categories not included in the framework. Consistent with current guidance for the field, we treated i-PARIHS as a set of proposed principles about how implementation works, and we interpreted each determinant in relation to that underlying account and to established implementation outcomes (32). The full coding framework is provided in Additional file 3.

Analysis followed the stages of the framework analysis method, namely familiarization, identifying a thematic framework, indexing, charting, and mapping and interpretation (30, 31, 33). The coding framework combined three elements. The first was a set of deductive codes corresponding to the four i-PARIHS constructs (18). The second was a set of study-specific deductive categories capturing PNCC practice, decision-making, attitudes toward guidelines, and program development. The third was a set of inductively derived codes added for salient concepts the existing framework did not represent including clinician autonomy and the standardization of practice. Throughout the analysis, we interpreted determinants in relation to the established implementation outcomes of acceptability, adoption, appropriateness, feasibility, fidelity, and sustainment which provided a shared vocabulary linking what participants described to outcomes the field aims to influence (32). All coding, indexing, and charting were conducted in NVivo 15 (Lumivero) (34).

Indexing proceeded in two phases. To develop and stabilize the framework, two coders independently indexed the same seven transcripts and then met to reconcile every code and to revise the codebook as needed. Where the coders could not agree on how a passage should be indexed, the first author adjudicated. This phase clarified code definitions and the relationship between the i-PARIHS and study-specific coding layers and produced the finalized codebook. The remaining 26 transcripts were then divided between the two coders and indexed independently against the finalized codebook, with the first author supervising and adjudicating queries throughout.

For the charting stage, indexed data were summarized into a single framework matrix in NVivo, with the i-PARIHS constructs and the inductively derived codes as columns and participants as rows, each classified by site and professional role. Each cell held a condensed summary, in the analysts’ words, of what a participant conveyed about a given construct. We then mapped and interpreted the charted data by examining patterns within each construct across participants and across rows to examine how constructs cohered within accounts, attending both to recurrent patterns and informative discrepant cases. The charted output is provided in Additional file 4. Sorting the matrix by site and role supported comparison across the three sites and across professional groups. All 33 transcripts were charted against the full framework, and each construct and category was checked for representation across the transcripts and across sites. Consistent with the study’s second aim, we used the charted matrix to assess how well each determinant was represented by i-PARIHS as currently specified to identify constructs for which the framework would benefit from elaboration and to map each candidate elaboration to the implementation outcomes it would be expected to influence.

The analytic team comprised the first author (M.T.V.), who conducted all interviews and supervised and adjudicated coding, and two coders (A.H. and A.J.), undergraduate students in public health trained in qualitative analysis. The coders were not affiliated with any participating institution and indexed from a position outside clinical practice which provided a check on clinically embedded assumptions. The team used ongoing discussion and memoing during indexing, charting, and interpretation. The 2025 participant-validation step provided a further check on the credibility of the findings.

Each i-PARIHS construct and study-specific category was evidenced across the transcripts, and every transcript contributed indexed data organized by the i-PARIHS constructs. Participants are identified by site and participant number (P1–P33); quotations are presented verbatim from transcripts of audio-recorded interviews. Determinants reported were identified at all three sites unless otherwise noted.

## Results

Thirty-three participants completed interviews: 11 at Site A, 10 at Site B, and 12 at Site C. Participants included pediatric intensivists, child neurologists, pediatric neurosurgeons, PICU advanced practice providers (nurse practitioners and advanced practice nurse managers), PICU bedside nurses, and physicians in departmental and hospital leadership roles. Many physician participants also held divisional, educational, or institutional leadership positions. At Site A, participants included six female and five male professionals; at Site B, five female, five male, and at Site C, six female, six male. The distribution of participants by professional role is shown in Table 1.

**Table 1.** Participants by site and professional role (N=33)

| Professional role | Site A | Site B | Site C | Total |
| --- | --- | --- | --- | --- |
| Physicians (intensivists, pediatric neurologists, neurosurgeons) | 6 | 8 | 10 | 24 |
| Nurse practitioners / advanced practice providers | 2 | 0 | 0 | 2 |
| Bedside / specialist nurses | 3 | 2 | 2 | 7 |
| Site total | 11 | 10 | 12 | 33 |

### The innovation, and how new practices were judged

Participants evaluated proposed practice changes against the strength of the supporting evidence, and several judged the legitimacy of a practice to its evidentiary basis. One neurologist mentioned that “every one of these protocols is backed up by a study” (Site B, P13).

However, evidence for a practice alone rarely determined uptake. A change was accepted more readily when its relative advantage over current practice was clear and when it fit existing workflows and values – as the innovation-side subconstructs i-PARIHS describes (18).

Participants weighed advantages against disadvantages explicitly, considering not only clinical benefit but cost and workload:

*Is there a cost? Is this going to cost the hospital system money? Is it going to lead to more or less consults and that could have an impact on the volume of work for a given consultant who maybe is already stretched too thin …* (Site C, P9)

Usability mattered. Practices that could be embedded where clinicians already worked, in the patient room or in the electronic medical record, were described as easier to adopt. This observation maps onto adoption and feasibility (18). Standardization of practice drew ambivalence, though it was valued for consistency and reducing unwarranted variation but resisted where clinicians felt it failed to accommodate the variability of individual patients. This tension recurs in the recipients and facilitation findings below.

### Recipients, and the capacity to absorb change

Receptivity to change was consistently described as a property of the recipients, and motivation, capacity, and time were discussed across all sites. Willingness to accept change was shaped by motivation, values and beliefs, prior experience, and the capacity to integrate a new practice into an already demanding workload. Motivation was often tied to outcomes; one intensivist described being “motivated and encouraged to try to meet those goals because the data has supported” improved outcomes (Site C, P14).

Values, however, could cut the other way, and participants contrasted receptivity to change with entrenched individual practice producing different responses depending on the recipient:

> *“There are people who are like, ‘This is the way it’s done because this is how I was trained to do it and this is my way.’”* (Site A, P6)

Collaboration, teamwork, and the influence of trusted local opinion leaders facilitated uptake, whereas limited time and competing clinical demands constrained it. Two determinants within this construct were emphasized by participants but are not named explicitly by i-PARIHS; we describe each in turn.

### Recipients: clinician autonomy (proposed)

A determinant that recurred across roles and sites, and that the original i-PARIHS framework does not name explicitly, was clinician autonomy, defined as the professional latitude to exercise independent judgment, long recognized as central to how clinicians experience their work and to how they receive external direction (35, 36). When asked whether guidelines affected their professional autonomy, most participants described guidelines as compatible with, or even supportive of, independent judgment rather than threatening to it. Autonomy functioned less as an obstacle to guidelines than as the lens through which their acceptability was judged. Changes framed as decision support that preserved judgment were acceptable, whereas changes perceived as mandates were not. One intensivist put it plainly:

> *Do they affect your professional autonomy? Not necessarily. I think … whenever I think about guidelines, I mean, as the name suggest[s] they’re guidelines, so you follow it. But if you want to deviate from those guidelines, you just need to have a reason why … And I think that’s absolutely fine as well*. (Site C, P24)

Others described guidelines as a starting point from which experienced clinicians could justifiably depart, a “base” from which to “launch practice” (Site A, P27), or as something that “enhances your critical thinking” rather than constraining it (Site A, P1).

The autonomy concern nonetheless surfaced. One nurse recalled a physician colleague who “felt offended” by guideline (Site C, P28).

Because the perceived threat to autonomy could operate as a barrier independent of the evidence for a change, and because it determined whether a change was judged acceptable and adopted, we treated clinician autonomy as a distinct concept within the recipient construct (Table 2, Proposition 1).

**Table 2.** i-PARIHS constructs and proposed elaborations in PNCC.

| i-PARIHS construct | What it captures here | What it does not capture | Proposed elaboration |
| --- | --- | --- | --- |
| Innovation | Evidence and knowledge sources; relative advantage; usability and workflow fit; observability. | Adequate as specified; ambivalence toward standardization is accommodated within fit and usability. | None required. |
| Recipients | Motivation tied to outcomes; values and beliefs; prior experience; capacity and time; opinion leaders. | Clinician autonomy as the lens for acceptability; the affective state clinicians bring to and take from change. | Add clinician autonomy and recipients' affective state as distinct determinants (Propositions 1 and 2). |
| Context | Team complexity and acuity; unit leadership and learning environment; organizational priorities and stability; evaluation structures. | That authority is distributed unequally across professions and that this distribution governs whether a change proceeds. | Re-specify the recipient subconstructs of power and authority and presence of boundaries as a team-level structural property, the interprofessional authority gradient (Proposition 3) |
| Facilitation | Dependence on credible champions and leadership; need for ongoing audit and re-education. | That facilitation must span professional boundaries and carry credibility across services to resolve disagreement. | Elaborate facilitation subconstructs to include authority-sensitive, boundary-spanning facilitation, consistent with prior PICU work. |

*i-PARIHS, integrated Promoting Action on Research Implementation in Health Services; PNCC,* *Pediatric Neurocritical Care; PICU, Pediatric Intensive Care Unit. For each construct, the table* *summarizes what i-PARIHS captured in PNCC, what it captured only partially, and the* *proposed elaboration. The recipients and facilitation elaborations build on the PICU i-PARIHS* *analysis of Steffen et al.: Steffen KM, Holdsworth LM, Ford MA, Lee GM, Asch SM, Proctor EK.* *Implementation of clinical practice changes in the PICU: a qualitative study using and refining* *the iPARIHS framework. Implement Sci. 2021;16(1):15. Propositions 1 to 3 are stated in the* *text.*

### Recipients: the affective state clinicians bring to change (proposed)

Participants across all three sites routinely described practice change and clinical decision-making in affective terms, including frustration, worry, the discomfort of uncertainty, moral distress, and the weight of high-stakes decisions. Bedside nurses in our study described the frustration of working between a standardized protocol and variable physician practice:

> *That … gets very frustrating. It’s very frustrating as a bedside nurse … knowing how to navigate that … It can get frustrating when we have this set protocol. We’re supposed to, you know, theoretically work through [it]*. (Site B, P29)

The emotional weight was voiced strongly by physicians. Participants described the frequent need for decision-making under uncertainty as stressful. Several also named moral distress explicitly, noting that deferring intervention could leave clinicians with the lingering “question … should we have done more,” which “can create some moral distress if […] things don’t go the way you expected” (Site A, P30).

Charting this determinant showed that emotional load shaped receptivity to change, since new protocols and service models could raise perceived risk, expose clinicians to moral conflict, set standardized expectations against individual judgment, and generate frustration when practice remained variable despite attempted standardization. That frustration bears directly on acceptability and sustainment because a change that is emotionally costly to maintain is less likely to persist (Table 2, Proposition 2).

### Context and interprofessional authority

The context of PNCC, namely large multiprofessional teams delivering high-stakes care, often at or near capacity, shaped every determinant. At the unit level, leadership support, a psychologically safe learning environment, and established mechanisms for embedding change facilitated practice change. Participants described disseminating changes through division meetings, in-person presentations, electronic order sets, and nursing “super users.” Receptivity also varied; one neurologist noted that those “who are aware of … recent developments” embraced change quickly, while others less comfortable with neurologic illness were slower to do so (Site C, P11).

At the organizational level, institutional priorities and senior leadership support determined whether resources followed, and instability could disrupt change. One participant described how a “complete administration flip” and the loss of a substantial share of PICU physicians made sustained change difficult (Site B, P5). Limited resources and infrastructure for evaluation and feedback constrained the ability to monitor whether changes achieved their intended effect.

### Context: an interprofessional authority gradient (proposed)

The determinant participants emphasized most strongly and the one that most clearly extended the i-PARIHS recipients and context constructs was an interprofessional authority gradient. PNCC is delivered by intensivists, neurologists, and neurosurgeons whose recommendations did not, in participants’ accounts, carry equal weight.

Physicians at all three sites described how the preferences of some services, frequently neurosurgery, could take precedence over others:

> *The preference of the neurosurgeons is oftentimes a priority and our preferences are overlooked.* (Site A, P6)

or

> *… they’re more likely to listen to the neurosurgeons than they are to us*. (Site C, P11)

In another example, recounting a disagreement over sedation, one intensivist observed that “at the end of the day, surgery had the final say” (Site C, P19), and another distinguished interventions they owned from those they did not, stating, “I cannot force a surgeon to go to the operating room” (Site A, P27).

Authority did not rest on professional title alone. One participant described a colleague who “doesn’t have positional power in the sense of having a title, but … has tons of authority because he is a wonderful human being to work with” (Site A, P10). Another reflected that “gender differences” could influence whether recommendations were followed (Site C, P11).

Participants were also alert to how implementing a change could itself “change the power dynamic” among services and roles (Site A, P22). i-PARIHS includes power and authority and presence of boundaries as recipient subconstructs; our data suggest further elaboration is needed to represent how authority is distributed across professions in a multidisciplinary setting, and how that distribution determines whether a given change, affecting adoption and fidelity, can proceed (Table 2, Proposition 3).

### Facilitation that was variable and under-resourced

Facilitation defined as the deliberate process of support and problem-solving through which the other constructs are activated (18), was described as variable and frequently under-resourced. Where a credible local champion or leader engaged colleagues and organized the work, change progressed; participants repeatedly identified leadership as decisive, with one observing that “you just need … like a kind of a point person” to organize and motivate colleagues (Site B, P25). In addition, dedicated administrative time and support are required to keep initiatives from stalling. Furthermore, participants emphasized the importance of collaboration across the services involved in PNCC; one physician leader stated plainly that the institution did “not have … adequate collaboration” between neurology, neurosurgery, and neurocritical care (Site A, P26).

Effective facilitation in this setting required someone credible across professional groups, able to convene disagreement, negotiate ownership of a change, and keep implementation from becoming a contest between services. We refer to this as *authority-sensitive facilitation*. A facilitator without standing across the relevant services could organize logistics but could not resolve the cross-professional disagreements on which adoption and sustainment depended.

Sustaining a change once introduced was described as requiring active, ongoing effort. One participant captured the principle as requiring continuous audit and feedback, “in God we trust; everyone else must bring data” (Site A, P31). The innovation, recipients, context, and facilitation constructs each captured a meaningful and distinct family of determinants. Table 2 summarizes what i-PARIHS captures for each construct, what it does not, and the proposed elaboration.

Table 3 then takes each elaboration from Table 2 a step further, linking it to the implementation outcomes it should affect and to the kind of strategy it implies.

**Table 3.**
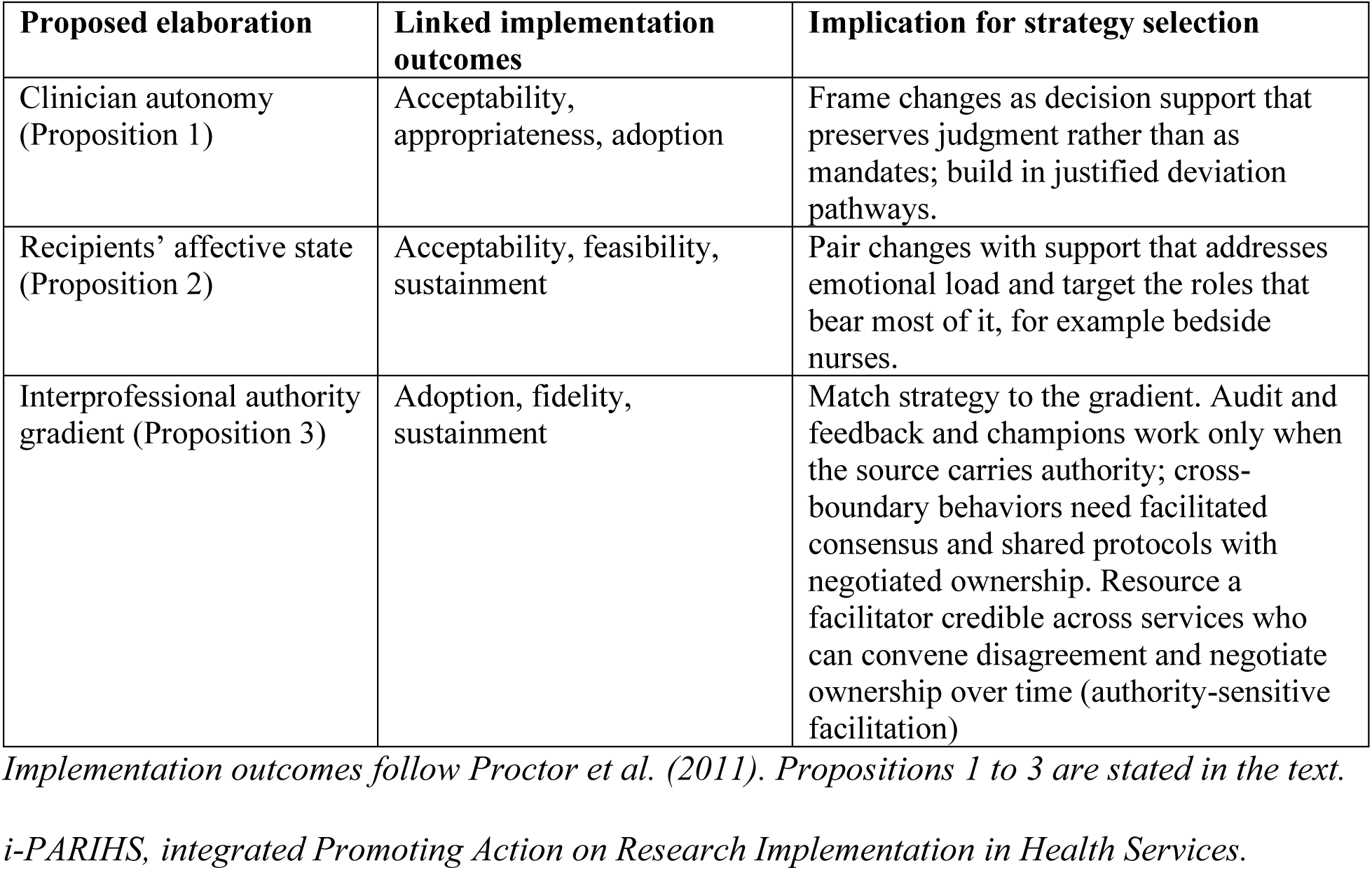
Proposed i-PARIHS elaborations, linked implementation outcomes, and strategy implications.

## Discussion

In this multi-site qualitative study, the determinants of practice change in PNCC were shaped less by the evidence base for a given change than by who was being asked to adopt a change, the multiprofessional environment in which they worked, and the quality of facilitation available to support the change. i-PARIHS organized these determinants well, but the data indicate three points at which the framework, as currently specified, only partially captured the PNCC environment. Here we have shown that in a high-acuity specialty where intensive care, neurology, and neurosurgery share patients and decisions, implementation is conditioned by professional discretion, affective load, and authority gradients that determine whether a change can be owned across services. These three determinants sit at the edges of innovation, recipients, context, and facilitation as i-PARIHS specifies them.

Clinician autonomy, within the recipients construct, is the professional latitude to exercise independent judgment. In our data, it governed how acceptability was judged; the same change was acceptable as decision support and unacceptable as a mandate. i-PARIHS names recipients’ motivation, values, and beliefs but does not name this latitude in relation to professional authority to exercise independent judgment as a determinant. The PICU i-PARIHS study noted loss of autonomy under the innovation construct, where it also affected recipients (22); our data locate it within the recipients construct, as the lens through which acceptability is judged. We propose adding it there, which yields a testable claim that a change framed as preserving clinician autonomy will be adopted more readily than an equivalent change framed as a mandate, independent of the strength of its supporting evidence (Proposition 1).

The affective dimension of recipients is the emotional state that clinicians bring to and take from a change, including worry, frustration, the discomfort of uncertainty, and moral distress. The emotional cost of working in critical care is well documented; moral distress and burnout are prevalent among ICU clinicians and closely linked, and nurses and other non-physician staff often report higher moral distress than physicians (37–41). The blood transfusion PICU i-PARIHS study also found providers’ emotions to be conceptually distinct from motivation, values, and beliefs, a distinction that made the affective dimension a candidate for addition to the framework (22). Other work has also argued that the emotional experience of those carrying out change shapes whether facilitation succeeds (42). Our findings add independent confirmation of this dimension, demonstrated most sharply by bedside nurses whose work was situated between a standardized protocol and variable physician practice. We propose that i-PARIHS represent recipients’ affective state explicitly, as a determinant distinct from values and beliefs, which yields a testable claim that facilitation addressing the emotional load of a change and attending to roles that bear most of it will sustain change more reliably than facilitation addressing knowledge and logistics alone (Proposition 2).

We define the interprofessional authority gradient as the unequal weight that different professions’ recommendations carry within a multidisciplinary team, such that the standing of the sponsoring profession of a proposed change helps determine whether the change proceeds. i-PARIHS includes power and authority and presence of boundaries among its recipient subconstructs and treats both as attributes of individual recipients. In our data, authority also functioned as a structural property of the team, one that distributed influence across professions. In PNCC, intensivists, neurologists, and neurosurgeons share patients but not equal standing, a pattern consistent with accounts of the ICU as a site of interprofessional negotiation in which collaboration follows implicit rules of the game and professional groups work to maintain their standing (43–45). The gradient also reaches the learning environment. Psychological safety in teams depends on leader inclusiveness and varies with professional status (45, 46), so the unit-level enablers participants described, leadership support and a psychologically safe learning environment, are unlikely to reach every profession on the team or be experienced by each team member in the same way. Participants’ observation that gender could influence whether recommendations were followed points in the same direction; concern about power and voice across the professions in healthcare teams is long-standing (47). We propose elaborating the power-and-authority and boundaries subconstructs to represent authority as a cross-professional gradient which yields a testable claim that in a multidisciplinary specialty, the position of a change’s sponsoring profession on that gradient predicts adoption over and above the change’s evidence, usability, and fit (Proposition 3).

Conceptualizing authority as a structural property of the team has implications for implementation strategies. Audit and feedback may not change behavior when the feedback originates from a profession whose recommendations carry less weight on the relevant gradient. A clinical champion may not change practice when the champion sits outside the dominant professional hierarchy for the behavior in question. When a target behavior crosses service boundaries, for example a sedation or neuromonitoring practice, strategies that build cross-service agreement, such as shared protocols with negotiated ownership (48), are more likely to succeed. Authority, in this case, conditions the mechanism through which a strategy is expected to work, and selecting strategies without accounting for it risks predictable failure. The same logic applies to autonomy and affect. A mandate-framed change can fail on acceptability however strong its evidence, and a change that is emotionally costly to maintain can fail on sustainment however feasible it first appears.

The challenge is familiar in quality improvement and implementation work where new practices are often only partially sustained (49). Consistent with prior work in the PICU (22), our data suggest that the facilitation construct would benefit from further elaboration of its subconstructs to capture how facilitation operates across professional boundaries (50) and over time (42), including the authority-sensitive facilitation described here.

These observations have practical implications for institutions developing PNCC programs. They suggest that implementation strategies attending only to the evidence for a practice change will be insufficient. Implementation efforts should additionally address recipients’ autonomy and the emotional experience of change by engaging the interprofessional authority structure explicitly. The need for credible, resourced facilitation is consistent both with the broader implementation science literature and with reports that successful PNCC program development rests on multidisciplinary collaboration (1, 8, 51). Detailed, practical guidance for program design will be reported in a companion paper.

This study makes three contributions. First, it corroborates, in an additional and more specialized setting, two elaborations that the PICU transfusion study had identified as candidates – clinician autonomy and the affective dimension of change (22). Determinant frameworks are widely applied and rarely revised once published, so corroboration of proposed refinements across settings has value in its own right. Second, it identifies a determinant absent from that earlier work, the interprofessional authority gradient, which participants traced to the tri-disciplinary interface among intensive care, neurology, and neurosurgery that characterizes neurocritical care. Third, it converts these observations into three testable propositions linked to implementation outcomes and strategy implications (Tables 2 and 3), and extends the earlier recommendation to elaborate facilitation (22) into authority-sensitive facilitation. A broader implication follows. Frameworks, like the interventions they help implement, may need tailoring to the setting in which they are applied.

## Strengths and limitations

Strengths of this study include its multi-site design, the inclusion of the full range of professional roles involved in PNCC, the use of an established implementation framework and analytic method, and the validation of preliminary findings with participants. Several limitations should be considered. A co-author led the earlier application of i-PARIHS in the general PICU with which two of our three determinants converge. That convergence therefore reflects a program of work extending across settings, not independent replication by an unrelated team. Interviews were conducted August 2023 to March 2024, and a 2025 participant-validation step supported the continued relevance of the findings and allowed additional reporting of new information. Practice may have evolved at individual sites, but the determinants reported here concern generally stable structural features of the care environment, namely team composition, professional roles and relationships, workflow demands, and decision-making under uncertainty. The three participating academic hospitals may not represent the full range of institutions delivering PNCC, including community and non-U.S. settings, which may limit generalizability. Findings reflect participants’ perspectives and were not triangulated against observational or quantitative data; mixed-methods extension is a direction for future work. The three propositions advanced here are derived from qualitative data and remain to be tested; doing so is the natural next step.

## Conclusions

The PNCC environment substantially shaped the determinants of practice change. i-PARIHS provided a useful structure for organizing those determinants and was adequate for identifying barriers and facilitators. When applied to this multidisciplinary specialty, the framework benefited from elaboration to represent clinician autonomy, the emotional dimension of change, and an interprofessional authority gradient, each linked here to the implementation outcomes and implications for strategy selection. As institutions organize PNCC into dedicated programs, attention to these determinants, and to credible, resourced, authority-sensitive facilitation, may improve the prospects for successful and sustained practice change.

## Supporting information

COREQ Checklist

Interview Guide

Codebook

Framework Analysis Table

## List of abbreviations

APP: advanced practice provider
COREQ: Consolidated Criteria for Reporting Qualitative Research
cEEG: continuous electroencephalography
EMR: electronic medical record
ICU: intensive care unit
i-PARIHS: integrated Promoting Action on Research Implementation in Health Services
PARIHS: Promoting Action on Research Implementation in Health Services
PICU: Pediatric Intensive Care Unit
PNCC: Pediatric Neurocritical Care
SRQR: Standards for Reporting Qualitative Research
TBI: traumatic brain injury.

## Declarations

### Ethics approval and consent to participate

The study was approved by the Institutional Review Board at University of California, Los Angeles (Study # 23-000248). Verbal informed consent was obtained from all participants prior to their interviews.

### Consent for publication

Not applicable. Quotations are presented in de-identified form, attributed by site and participant number only.

### Availability of data and materials

Given the risk of re-identification in a small multiprofessional sample, the full interview transcripts are not publicly available. De-identified excerpts may be available from the corresponding author on reasonable request, subject to the governing ethics approval.

### Authors’ contributions

M.T.V. wrote the original draft and contributed to formal analysis, validation, resources, investigation, project administration, visualization, and supervision. B.D.S., J.S.W., and J.A.P. led participant recruitment as part of investigation. M.T.V., B.D.S., J.S.W., P.C., and J.A.P. contributed to validation. A.H. and A.J. contributed to formal analysis supervised by M.T.V. K.M.S. contributed to the conceptualization and methodology of the parent framework analysis and to review and editing. J.A.P. contributed funding acquisition and supervision. All authors read, reviewed, edited, and approved the final manuscript.

## Acknowledgements

The authors thank the providers at the participating institutions who gave their time to be interviewed.

## Additional files

Additional file 1. Additional_file_1_COREQ_checklist.docx. Consolidated Criteria for Reporting Qualitative Research checklist. Completed 32-item COREQ checklist indicating where each item is reported in the manuscript.

Additional file 2. Additional_file_2_Interview_guide.docx. Semi-structured interview guide. Full guide used in all 33 interviews, including the oral consent script, the four topic sections, and the role-specific modifications for nurses, with identifying references replaced by placeholders.

Additional file 3. Additional_file_3_Coding_framework.docx. Coding framework. Deductive i-PARIHS constructs and subconstructs with definitions, study-specific deductive categories, and inductively derived codes, as applied in NVivo 15.

Additional file 4. Additional_file_4_Framework_analysis_table.docx. Description of themes and sub-themes with examples. Charted output of the framework analysis, mapping i-PARIHS constructs and subconstructs to the themes and sub-themes developed during interpretation, with descriptions and illustrative quotations.

