## Supplementary material for "Innovations, recipients, and contexts in the implementation of pediatric neurocritical care programs: a multisite qualitative study refining i-PARIHS": COREQ Checklist

### Additional file 1. COREQ checklist

*Consolidated Criteria for Reporting Qualitative Research (COREQ), 32-item checklist. Locations refer to sections of the manuscript. Page numbers should be added after final typesetting.*

| No. | Item | Guide question / description | Reported in manuscript |
|---|---|---|---|
| Domain 1: Research team and reflexivity | | | |
| 1 | Interviewer / facilitator | Which author conducted the interview or focus group? | Methods, Setting and participants; Analysis. All interviews were conducted by the first author (M.T.V.). |
| 2 | Credentials | What were the researcher's credentials? | Title page. M.T.V. holds a PhD, MPH, and MSW. |
| 3 | Occupation | What was their occupation at the time of the study? | Title page; Methods, Setting and participants. Public health researcher (PhD, MPH, MSW) with a disciplinary background in public health, social work, and implementation science. |
| 4 | Gender | Was the researcher male or female? | Declarations, Competing interests. The interviewer is female. |
| 5 | Experience and training | What experience or training did the researcher have? | Methods, Setting and participants. Interviews were conducted by a team member with qualitative research experience whose disciplinary background is in public health, social work, and implementation science. |
| 6 | Relationship established | Was a relationship established prior to study commencement? | Methods, Setting and participants. A site representative assisted with identifying and recruiting eligible providers. No prior relationship between the interviewer and participants is reported. |
| 7 | Participant knowledge of the interviewer | What did participants know about the researcher? | Methods, Setting and participants; Additional file 2. The consent script identified the interviewer and stated that findings would inform PNCC program development at the participant's institution. |
| 8 | Interviewer characteristics | What characteristics were reported about the interviewer? | Methods, Setting and participants. The interviewer’s qualitative research experience and disciplinary background are reported. |
| Domain 2: Study design | | | |
| 9 | Methodological orientation and theory | What methodological orientation was stated to underpin the study? | Methods, Study design and reporting; Analysis. Qualitative interview study using framework analysis, with i-PARIHS as the guiding determinant framework. |
| 10 | Sampling | How were participants selected? | Methods, Setting and participants. Purposive sampling across professional roles involved in PNCC at three sites. |
| 11 | Method of approach | How were participants approached? | Methods, Setting and participants. Participants were identified and recruited with the assistance of a site representative. |

| No. | Item | Guide question / description | Reported in manuscript |
|---|---|---|---|
| 12 | Sample size | How many participants were in the study? | Results. Thirty-three providers completed interviews, 11 at Site A, 10 at Site B, and 12 at Site C. |
| 13 | Non-participation | How many people refused to participate or dropped out, and why? | Not reported. |
| 14 | Setting of data collection | Where was the data collected? | Methods, Setting and participants. Interviews were conducted remotely using a video-conferencing platform. |
| 15 | Presence of non-participants | Was anyone else present besides the participants and researchers? | Not reported. |
| 16 | Description of sample | What are the important characteristics of the sample? | Results and Table 1. Professional role, site, and gender distribution are reported. |
| 17 | Interview guide | Were questions, prompts, or guides provided by the authors? Was it pilot tested? | Methods, Interview guide; Additional file 2. The full guide is provided. Pilot testing is not reported. |
| 18 | Repeat interviews | Were repeat interviews carried out? | No. Each participant was interviewed once. |
| 19 | Audio / visual recording | Did the research use audio or visual recording to collect the data? | Methods, Setting and participants. Interviews were audio-recorded and transcribed verbatim. |
| 20 | Field notes | Were field notes made during and/or after the interview or focus group? | Methods, Analysis. The team used ongoing memoing during indexing, charting, and interpretation. Field notes taken during interviews are not reported. |
| 21 | Duration | What was the duration of the interviews or focus group? | Methods, Data collection period and participant validation. Interviews lasted approximately 25 to 85 minutes (site medians 53 to 59 minutes). |
| 22 | Data saturation | Was data saturation discussed? | Methods, Setting and participants. Sampling continued until the team judged that further interviews were unlikely to alter the determinant structure. |
| 23 | Transcripts returned | Were transcripts returned to participants for comment and/or correction? | No. Transcripts were not returned. Preliminary site-level findings were shared with participants for validation (see item 28). |
| Domain 3: Analysis and findings | | | |
| 24 | Number of data coders | How many data coders coded the data? | Methods, Analysis. Two coders (A.H., A.J.) indexed the data, with the first author supervising and adjudicating. |
| 25 | Description of the coding tree | Did authors provide a description of the coding tree? | Methods, Analysis; Additional file 3. The full coding framework is provided. |
| 26 | Derivation of themes | Were themes identified in advance or derived from the data? | Methods, Analysis. Deductive i-PARIHS codes and study-specific categories were specified in advance; additional codes were derived inductively. |

| No. | Item | Guide question / description | Reported in manuscript |
|---|---|---|---|
| 27 | Software | What software, if applicable, was used to manage the data? | Methods, Analysis. NVivo 15 (Lumivero). |
| 28 | Participant checking | Did participants provide feedback on the findings? | Methods, Data collection period and participant validation. Preliminary site-level findings were shared with participants in October 2025 for validation. |
| 29 | Quotations presented | Were participant quotations presented to illustrate the themes or findings? | Methods, Analysis; Results. Quotations are presented verbatim and identified by site and participant number. |
| 30 | Data and findings consistent | Was there consistency between the data presented and the findings? | Results and Discussion. Each reported determinant is supported by quoted data. |
| 31 | Clarity of major themes | Were major themes clearly presented in the findings? | Results. Findings are organized under the i-PARIHS constructs with subheadings. |
| 32 | Clarity of minor themes | Is there a description of diverse cases or discussion of minor themes? | Methods, Analysis; Results. Discrepant cases were examined during mapping and interpretation and are reported alongside recurrent patterns. |

Reference: Tong A, Sainsbury P, Craig J. Consolidated criteria for reporting qualitative research (COREQ): a 32-item checklist for interviews and focus groups. Int J Qual Health Care. 2007;19(6):349-57.
