## Supplementary material for "Innovations, recipients, and contexts in the implementation of pediatric neurocritical care programs: a multisite qualitative study refining i-PARIHS": Interview Guide

### Additional file 2. Interview guide

*Semi-structured interview guide used in all 33 interviews, September 2023 to March 2024. The guide was developed from the i-PARIHS framework. Identifying references to the interviewer's institution, the participating sites, and the reviewing institutional review board have been replaced with bracketed placeholders for de-identification. Content is otherwise reproduced as used.*

#### Introduction and consent

##### Oral consent script

Hello, this is [interviewer] from [institution]. We had set up this time to talk to you about your perspective on pediatric neurocritical care. Is this still a good time to talk?

[If no, try to schedule another time. If yes, keep reading.]

Great. I'll read you the consent script.

Participation is voluntary.

The interview will last approximately 60 minutes and will be recorded on Zoom. The degree of risk is perceived as very minimal in this study, although you may be uncomfortable when asked certain questions. You are not required to answer the questions and may end the interview at any time.

Results from this research will help assess current practices, readiness for change, and inform our development of a Pediatric Neurocritical Care Program at your institution.

Recordings will be destroyed once contents have been transcribed. Your name will not be linked with your answers.

Your de-identified data may be kept for use in future research.

If you have questions about your rights as a research subject, you may call [the reviewing institution's human research protection program] at [telephone] or via email at [email address]. Would you like me to repeat the contact information?

Do you have any questions about this research?

Do you consent to participating in this interview and being recorded?

[If yes, continue with interview. If no, thank them for their time.]

Great. Let's get started. I am now going to turn on the recorder.

[Goal: applying the Promoting Action on Research Implementation in Health Services (i-PARIHS) implementation science framework to assess practice and willingness to accept changes in practice.]

[Deliverable: knowledge that will inform our implementation planning and strategy selection for the development of a Pediatric Neurocritical Care Program.]

#### Section 1. Change and impact of change in the ICU

*The first section focuses on general questions about how change occurs in your ICU as it relates to pediatric neurocritical care practices. Are you ready to get started?*

**0. What is your role at the hospital?**

**1. What is the working environment like as it relates to changes in neurocritical care practices?**

- Who usually has a say in terms of change, as it relates to pediatric neurocritical care?
- What are the usual barriers to change?
- What types of things help change occur?
- Do you feel like you have a role or ownership in any changes that occur, as it relates to pediatric neurocritical care?
- When it comes to pediatric neurocritical care at your institution, what makes change successful or unsuccessful?
- What makes change stick, or be sustainable in your unit?
- Does the hospital facilitate change or innovation in the neurosciences, including neurocritical care? How?
- Tell me about a change in pediatric neurocritical care practice that has recently occurred.
- Who proposed the change?
- How was this proposal made?
- Who were the thought leaders who drove the change?
- How did you find out about the change? What are the most effective ways people find out about change in your unit?
- How did this change occur in your unit, for example, who enacted the change?
- What methods were used to facilitate change?
- What was your role in the change?
- What went well with this change?
- How could it have been improved?

**2. How do you typically respond to changes in neurocritical care practices?**

- What makes being receptive to change easier?
- What makes being receptive to change more difficult?
- How do you see others respond to changes in neurocritical care practices?
- Who is accountable for change?
- How do we track change, and do we have the necessary tools or resources?
- Could you share a change in neurocritical care practice that you were, or would be, receptive to, and an example of a change you were not or would not be receptive to?

**3. If you had a concern or problem related to delivery of neurocritical care, who would you discuss it with? For example, peers, leadership, or administration?**

- To what extent do you feel supported?
- Who do others outside your role or group depend on?

**4. When it comes to neurocritical care patients, how do you know you've done a good job?**

- How do you find out if there are areas where you need to improve?
- How do others get feedback on their work?

**5. Is there anything I haven't asked about that would be important for me to understand about the working environment in the ICU as it relates to changes in neurocritical care practices?**

#### Section 2. Neurocritical care decision making and attitudes toward a neurocritical care program

**6. What is your understanding of the current state of neurocritical care practices in your PICU? For example, would you say there is a programmatic approach to it? Is it targeted and organized, with specific goals, metrics for success, and resources for neurocritical care? If not, what components of a programmatic approach to neurocritical care would be highest priority or most valuable?**

**7. Can you identify anticipated barriers to the implementation of a neurocritical care program, from within the ICU to the healthcare system level?**

*The upcoming questions relate to the care of critically ill children with acute neurological or neurosurgical conditions, for example cerebral hemorrhage, ischemic stroke, traumatic brain injury, seizures, and post-operative care after brain tumor or scoliosis surgery.*

**8. Please describe how you decide to intervene when caring for children with acute neurological injury or clinical changes.**

*Nurse modification: From your point of view, what factors seem to influence providers in your unit to intervene when caring for children with acute neurological injury or clinical changes?*

- When considering physiologic parameters, do you use specific thresholds or physiological trends?
- Do you currently incorporate neurocritical care guidelines into your decision making? If yes, how frequently?
- Is your risk versus benefit assessment different for neurocritical care patients than for other patients?
- Are there disadvantages particular to neurocritical care interventions?
- Are there barriers to executing neurocritical care interventions you consider beneficial to patients, for example emergent neuroimaging or surgical interventions?

**9. How, if at all, has your approach to neurocritical care interventions changed over time?**

*Nurse modification: From your perspective, have you noticed changes over time in how or when people apply neurocritical care interventions?*

- Intervene more or less?
- Why?
- Can you share a couple of examples of studies that in your opinion guide neurocritical care interventions?
- What are your general thoughts about these studies, for example whether they are applicable to practice or of good quality?

**10. How does decision making flow regarding neurocritical care interventions in your unit?**

- Is decision making largely directed by attendings? If so, from which specialties?
- Is the approach to decision making different depending on those involved? How?
- How, if at all, has the approach to decision making for neurocritical care interventions changed over time?

**11. Do subspecialty groups other than neurology and neurosurgery influence which or when patients receive neurocritical care interventions?**

- In what situations?
- How are disagreements around neurocritical care interventions resolved in such situations?

**12. Can you identify barriers or facilitators in the process of implementing a neurocritical care intervention for the patient once you or someone else has decided to do it?**

- Process of ordering, administering, and tracking response.

**13. In what way is managing neurocritical care patients affected by what is going on in the ICU?**

- Whether the unit is busy in general?
- Whether you need to attend to a more urgent clinical issue?
- Whether any of the specialty teams caring for the patient are busy at the time, in or outside of the ICU?
- Anything else you can think of?

#### Section 3. Gray areas, watching and waiting

*This next section of questions is about how you might manage a neurocritical care patient with a controversial clinical scenario, when the decision about intervening might be difficult and there is a bit of a gray area.*

**14. What would you consider gray areas in neurocritical care?**

**15. What thought processes guide your decision to watch and wait instead of intervening when a decision is controversial?**

*Nurse modification: what thought processes guide the decision to watch and wait?*

- What influences you to watch and wait?
- How do other clinicians, medical staff including nurses and nurse practitioners, residents, fellows, and families impact your decision to watch and wait?
- Are there benefits to self, patients, colleagues, or the healthcare organization?
- Are there disadvantages?
- Does watching and waiting ever conflict with some other goal or objective in terms of patient care?
- How, if at all, does watching and waiting influence your work stress?

**16. Are there system changes or resources you can think of that would help address topics or challenges you have shared so far?**

#### Section 4. Review of neurocritical care guideline repository and contextual factors around implementation

**17. What are your thoughts about using guidelines when you care for neurocritical care patients?**

- Do you currently, or would you be willing to, use neurocritical care guidelines? Why?
- Are others in your unit willing to use such guidelines?
- Do you find them useful, or constraining? Do they affect your professional autonomy?

**18. Would neurocritical care guidelines change your approach to decision making for neurocritical care patients? How?**

*Nurse modification: How do you think guidelines would impact providers' approach to neurocritical care interventions?*

- Are there specific advantages to using guidelines when caring for neurocritical care patients?
- Are there disadvantages?
- What are the logistical challenges in your unit, or features of the guidelines themselves, that create barriers to using them?

*Share overview of current guidelines.*

**19. Have you seen these guidelines before? [Share by screen for remote interviews.]**

**20. Are you familiar with where they live and how to access them?**

**21. How important are these guidelines? How do these guidelines impact patients?**

- How well do these guidelines fit with your current practice and with current practice in your unit?
- What adaptations in your workflow or the guidelines themselves would help incorporate them better into practice?
- When in your workflow is the best time to present this kind of information to you?
- Assuming it was feasible, what would be the best way to make the guidelines available to you and other team members in your unit?
- Who would need to support the guidelines for them to be used more consistently, for example other subspecialists or information technology?

**22. What else would be important for me to understand about using guidelines?**

**23. What are your additional thoughts, recommendations, or concerns regarding the development and implementation of a pediatric neurocritical care program?**

**24. What would your vision of a pediatric neurocritical care program be at your institution?**
