## Supplementary material for "Innovations, recipients, and contexts in the implementation of pediatric neurocritical care programs: a multisite qualitative study refining i-PARIHS": Codebook

### Additional file 3. Coding framework

*Coding framework used for indexing all 33 transcripts in NVivo 15. The framework combined three elements: deductive codes corresponding to the i-PARIHS constructs and subconstructs; study-specific deductive categories developed for this study; and inductively derived codes added for concepts the existing framework did not represent. Two subconstructs marked below were added to the recipients construct by the authors during framework development.*

#### Part A. i-PARIHS constructs and subconstructs

| Construct | Subconstruct | Definition |
|---|---|---|
| Innovation | Underlying knowledge sources | Specified sources of information relevant to the specific innovation, including research or published guidelines, clinical and patient experience, and local practice information (1) |
| | Clarity | How well the innovation is understood by the recipients |
| | Degree of fit with existing practice and values | How an evidence-based practice aligns with individuals' or a group's own norms, values, and perceived risks and needs, and how it fits with existing workflows and systems (1, 2) |
| | Usability | The extent to which an innovation is able to be used in the work system |
| | Relative advantage | Stakeholders' perception of the advantage or disadvantage of implementing the evidence-based practice versus an alternative solution, or stakeholders' cost-benefit perceptions compared with other alternatives (1, 2) |
| | Trialability | The ability to test the evidence-based practice on a small scale in the organization, and to reverse course if warranted (1, 2) |
| | Observable results | The degree to which the results of an innovation are visible or able to be seen being used in practice by adopters (1, 2, 3) |
| Recipients | Motivation | The reasons one has for acting or behaving in a particular way, or the general desire or willingness of someone to do something |
| | Values and beliefs | Individual or group principles or standards of behavior, ideas deemed true, or what is determined to be important (3, 4) |
| | Goals | The end state or outcomes toward which an individual or group is striving (4) |
| | Skills and knowledge | Abilities or proficiencies obtained through training or practice, and the awareness of specific facts or concepts (4) |
| | Time, resources, support | Things or human resources used to achieve a goal, and anything that aids or assists in achieving a goal (4) |
| | Local opinion leaders | Individuals who are seen as likable, trustworthy, and influential, and who may be able to persuade others to implement a new innovation through their influence (5) |
| | Collaboration and teamwork | Cooperative effort toward a common goal or on a common project (4) |
| | Existing networks | The nature and quality of webs of social networks within an organization (2) |
| | Power and authority | The capacity or power to influence events, behaviors, situations, or people, even when others try to resist this influence (4) |
| | Presence of boundaries | The presence of barriers to sharing knowledge and spreading work practices (6) |

| Construct | Subconstruct | Definition |
|---|---|---|
| | Expectations or requirements (added to i-PARIHS by the authors) | Tasks and engagement or participation in efforts that people are expected or required to complete for their job. Applied when there is a disconnect between individuals or groups that vary in their ideas of what is expected or required |
| | Feelings or emotions (added to i-PARIHS by the authors) | Any expressed feeling or emotion a participant has about something related to work, implementation efforts, or the innovation itself |
| Context, local level (ICU level) | Formal and informal leadership support | The behaviors, attitudes, and actions of leaders that reflect readiness or receptivity to a change (1) |
| | Culture | Prevailing norms, values, and basic assumptions of a given organization, staff characteristics, or the way things are done here (1, 2) |
| | Past experience of innovation and change | Prior experience with the introduction of something new |
| | Mechanisms for embedding change | The process of incorporating a new thing or process such that it is fully implemented and consistently used in day-to-day work |
| | Evaluation and feedback processes | The systems and processes for collecting, displaying, and using data related to evidence-based practice in meaningful forms to measure practice or change (1) |
| | Learning environment | A climate in which leaders express their own fallibility and need for team members' assistance and input; team members feel they are essential, valued, and knowledgeable partners in the change process; individuals feel psychologically safe to try new methods; and there is sufficient time and space for reflective thinking and evaluation (2) |
| Context, organizational level (outside the ICU) | Organizational priorities | Values and goals that guide the actions of an organization |
| | Senior leadership and management support | The behaviors, attitudes, and actions of hospital leaders outside and above the ICU that reflect readiness or receptivity to a change |
| | Culture | Prevailing norms, values, and basic assumptions of a given organization, or the way things are done here (1, 2) |
| | Structure and systems | The social architecture, age, maturity, and size of an organization (2) |
| | History of innovation and change | Prior experience with the introduction of something new |
| | Absorptive capacity | The capacity for change, preparedness, and receptivity of involved individuals to an innovation, and the extent to which use of that innovation will be rewarded, supported, and expected within their organization (1, 2) |
| | Learning networks | Continuing education, collaboratives, or other organized efforts to enhance an individual's or group's knowledge |
| Facilitation | Facilitation | A deliberate process of interactive problem solving and support that occurs in the context of a recognized need for improvement and a supportive interpersonal relationship (1) |

#### Part B. Inductively derived codes

| Code | Definition |
|---|---|
| Consistent exposure to intervention | The idea that implementation is easier or more sustainable when individuals can use the intervention frequently |
| Provider autonomy | Self-directed and independent activity by a care provider, for example in decision making, usually in the context of a provider discussing the value of autonomy or fear of losing autonomy |
| Response to bad outcome | Quality or improvement efforts that occur in response to events where a negative outcome occurred |
| Standardization of practice | Ideas or opinions, favorable or unfavorable, relating to consistency of medical practice between patients or over time |

#### Part C. Study-specific deductive categories

| Category | Subcategory | Definition |
|---|---|---|
| Current neurocritical care environment and practices | Role and responsibilities | Descriptions of the participant's role in neurocritical care practice and decision making |
| | Existing PNCC practices | Overview of current neurocritical care procedures, pathways, and informal norms |
| Organizational climate for change | Change culture | Descriptions of how change generally occurs within the PICU environment |
| | Barriers to change | Challenges or resistance faced in implementing change |
| | Facilitators of change | Enablers or supports that help change happen successfully |
| | Ownership and accountability | Descriptions of individual or group ownership of change |
| | General suggestions | General suggestions offered by participants |
| | Suggestions for sustainability | Suggestions for changes that might not be part of the PNCC program specifically |
| Decision making in neurocritical care | Who do you talk to about concerns in PNCC | Applied when participants speak about who they go to for help or to express concerns about care. Consider also coding power and authority |
| | Decision making processes | How decisions are made in neurocritical care cases |
| | Influence of subspecialties | Influence of teams beyond or including neurology and neurosurgery |
| | Watching and waiting | Experiences with clinical uncertainty and delayed action |
| | Use of technology and physiological trends | Use of technology and physiological trends in the decision making process |
| Sense of support | Sense of support | The extent to which participants feel supported |
| Receptivity to change | Personal response to change | The participant's own reactions to changes in PNCC practice |
| | Unit-level response to change | Observations of team or peer responses to changes |

| Category | Subcategory | Definition |
| --- | --- | --- |
| Feedback and performance assessment | Evaluating clinical performance | How individuals and teams assess effectiveness of PNCC care |
| Attitudes toward guidelines | Perceived value of guidelines | Beliefs about usefulness of PNCC guidelines |
| | Barriers to guideline use | Reasons guidelines are not used in practice |
| | Guideline integration into workflow | Suggestions for making guideline use feasible |
| | How providers access the guidelines | How providers access the guidelines. Consider also coding from the innovation codes as appropriate |
| | Knowledge and awareness of the guideline | Knowledge of the guideline and where it exists. Consider also coding from the innovation codes as appropriate |
| Barriers and facilitators to interventions | Barriers (clinical) | Challenges in executing neurocritical care interventions |
| | Facilitators (clinical) | Supports for enacting neurocritical care interventions |
| Programmatic vision for PNCC | Ideal features of a PNCC program | What should be included in a new program, in an ideal world |
| | Anticipated barriers to a PNCC program | Predicted barriers to starting a PNCC program |
| | Recommendations for program development | Concrete suggestions for developing the program, including protocols that should be developed |
