## Supplementary material for "Innovations, recipients, and contexts in the implementation of pediatric neurocritical care programs: a multisite qualitative study refining i-PARIHS": Framework Analysis Table

**Additional file 4** Description of themes and sub-themes with examples

*Charted output of the framework analysis. Each row records an i-PARIHS construct and subconstruct against which data were indexed, the theme and sub-theme developed during mapping and interpretation, a description of the charted content, and an illustrative quotation. Rows marked as a proposed elaboration identify determinants that the framework, as currently specified, captured only partially. Quotations are attributed by de-identified site and participant number.*

| i-PARIHS construct(s) | i-PARIHS subconstruct(s) | Theme | Sub-theme | Description | Example |
|---|---|---|---|---|---|
| Recipients; Context | Values and beliefs; Motivation; Local level: Culture | How new practices were judged | Evidence necessary but not sufficient | Participants evaluated proposed changes against the strength of the supporting evidence, but evidence alone rarely determined uptake. A change was accepted more readily when its relative advantage over current practice was clear and when it fit existing workflows and values. | "every one of these protocols is backed up by a study" (Site B, P13) |
| Innovation | Relative advantage; Degree of fit with existing practice and values | How new practices were judged | Cost and workload weighed explicitly | Participants weighed advantages against disadvantages explicitly, considering not only clinical benefit but cost and workload. | "Is there a cost? Is this going to cost the hospital system money? Is it going to lead to more or less consults and that could have an impact on the volume" (Site C, P9) |
| Innovation | Usability; Observable results | How new practices were judged | Embedding where clinicians already work | Practices that could be embedded in the patient room or in the electronic medical record were described as easier to adopt, and participants valued seeing a change in use before fully committing to it. Maps onto adoption and feasibility. | "tied into the electronic medical record, tied into workflows. Minimizing the amount of remembering that you have to do" (Site B, P33) |
| Innovation; Recipients | Degree of fit with existing practice and values; Values and beliefs | How new practices were judged | Ambivalence toward standardization | Standardization of practice, indexed inductively, was valued for consistency and for reducing unwarranted variation, but resisted where clinicians felt it failed to accommodate the variability of individual patients. | "guidelines allow us to find the patient who's the exception ... but that the majority of patients are not the exception and when you standardize care[,] care is better" (Site A, P10) |
| Recipients | Motivation; Values and beliefs; Skills and knowledge; Time, resources, support | Capacity to absorb change | Receptivity as a property of the recipient | Willingness to accept change was shaped by motivation, values and beliefs, prior experience, and the capacity to integrate a new practice into an already demanding workload. The same innovation produced different responses depending on the recipient. | "motivated and encouraged to try to meet those goals because the data has supported" improved outcomes (Site C, P14) |

| | | | | | |
|---|---|---|---|---|---|
| Recipients | Values and beliefs; Past experience of innovation and change | Capacity to absorb change | Entrenched individual practice | Values could cut the other way. Participants contrasted a pro-change disposition with entrenched individual practice. | "There are people who are like, 'This is the way it's done because this is how I was trained to do it and this is my way." (Site A, P6) |
| Recipients | Collaboration and teamwork; Local opinion leaders; Time, resources, support | Capacity to absorb change | Teamwork and opinion leaders against limited time | Collaboration, teamwork, and the influence of trusted local opinion leaders facilitated uptake, whereas limited time and competing clinical demands constrained it. | "when the team works well together ... it's like a well-oiled machine and everything happens the way it's supposed to" (Site B, P13); "just the bandwidth of the number of things that need to get accomplished within a whole unit" (Site B, P15) |
| Recipients (proposed elaboration) | Values and beliefs; Motivation (autonomy not named explicitly) | Clinician autonomy | Autonomy as the lens for judging acceptability | Clinician autonomy, the professional latitude to exercise independent judgment, recurred across roles and sites. Most participants described guidelines as compatible with, or supportive of, independent judgment rather than threatening to it. Autonomy functioned less as an obstacle than as the lens through which acceptability was judged. | "Do they affect your professional autonomy? Not necessarily. I think ... whenever I think about guidelines, I mean, as the name suggest[s] they're guidelines" (Site C, P24) |
| Recipients (proposed elaboration) | Values and beliefs; Skills and knowledge | Clinician autonomy | Guidelines as a starting point | Experienced clinicians described guidelines as a base from which practice could justifiably depart, rather than as a constraint. | "base" from which to "launch practice" (Site A, P27) |
| Recipients (proposed elaboration) | Values and beliefs; Feelings or emotions | Clinician autonomy | Perceived threat to autonomy as a barrier | The autonomy concern surfaced directly where a change was experienced as a judgment on competence. The perceived threat could operate as a barrier independent of the evidence for a change, and determined whether a change was judged acceptable and adopted. | a physician colleague "felt offended" by guidelines (Site C, P28) |
| Recipients (proposed elaboration) | Feelings or emotions (added to i-PARIHS by the authors) | Affective state clinicians bring to change | Frustration between protocol and variable practice | Bedside nurses described the frustration of working between a standardized protocol and variable physician practice. | "That ... gets very frustrating. It's very frustrating as a bedside nurse ... knowing how to navigate that" (Site B, P29) |

| | | | | | |
|---|---|---|---|---|---|
| Recipients (proposed elaboration) | Feelings or emotions; Motivation | Affective state clinicians bring to change | Emotional weight of high-stakes decisions | Participants described practice change and decision making in affective terms including frustration, worry, the discomfort of uncertainty, moral distress, and the weight of high-stakes decisions. Emotional load shaped receptivity, because a change that is emotionally costly to maintain is less likely to persist. Bears on acceptability and sustainment. | "watch and wait to see where the child is going to end up, that is stressful. It's particularly stressful ... on our nursing colleagues who literally are at the bedside for that twelve-hour shift and feel helpless" (Site A, P21) |
| Context | Local level: Formal and informal leadership support; Learning environment; Mechanisms for embedding change | Context of high-stakes multiprofessional care | Leadership, psychological safety, and channels for embedding change | At the unit level, leadership support, a psychologically safe learning environment, and established mechanisms for embedding change facilitated practice change. Changes were disseminated through division meetings, in-person presentation, electronic order sets, and nursing super users. | those "who are aware of ... recent developments" embraced change quickly (Site C, P11) |
| Context | Organizational level: Organizational priorities; Senior leadership and management support; Structure and systems | Context of high-stakes multiprofessional care | Organizational instability disrupts change | Institutional priorities and senior leadership support determined whether resources followed, and instability could disrupt change. Limited structures for evaluation and feedback constrained the ability to monitor whether changes achieved their intended effect. | a "complete administration flip" and the loss of a substantial share of PICU physicians made sustained change difficult (Site B, P5) |
| Recipients; Context (proposed elaboration) | Power and authority; Presence of boundaries | Interprofessional authority gradient | Unequal weight of service recommendations | PNCC is delivered by intensivists, neurologists, and neurosurgeons whose recommendations did not, in participants' accounts, carry equal weight. Physicians at all three sites described how the preferences of some services, frequently neurosurgery, could take precedence over others. | "The preference of the neurosurgeons is oftentimes a priority and our preferences are overlooked." (Site A, P6) |
| Recipients; Context (proposed elaboration) | Power and authority; Presence of boundaries | Interprofessional authority gradient | Consequential at the level of the individual case | The gradient determined whose preference prevailed in disagreements over management of individual patients. | "... they're more likely to listen to the neurosurgeons than they are to us." (Site C, P11) |

| | | | | | |
|---|---|---|---|---|---|
| Recipients (proposed elaboration) | Power and authority; Local opinion leaders | Interprofessional authority gradient | Authority resting on credibility rather than title | Authority did not rest on position alone. Influence was located in credibility rather than formal designation, and participants observed that gender differences could influence whether recommendations were followed. | a colleague who "doesn't have positional power in the sense of having a title, but ... has tons of authority because he is a wonderful human being to work with" (Site A, P10) |
| Recipients; Context (proposed elaboration) | Power and authority; Presence of boundaries | Interprofessional authority gradient | Change itself redistributes authority | Participants were alert to how implementing a change could itself alter the distribution of influence among services and roles. Affects adoption and fidelity. | implementing a change could "change the power dynamic" among services and roles (Site A, P22) |
| Facilitation | Facilitation | Variable and under-resourced facilitation | A credible point person | Where a credible local champion or leader engaged colleagues and organized the work, change progressed. Participants repeatedly identified leadership as decisive. | "you just need ... like a kind of a point person" (Site B, P25) |
| Facilitation; Context | Facilitation; Presence of boundaries | Variable and under-resourced facilitation | Facilitation without time or support stalls | Where facilitation depended on individual initiative without dedicated time or support, it stalled. Participants described limited collaboration across the services involved in PNCC. | the institution did "not have ... adequate collaboration" between neurology, neurosurgery, and neurocritical care (Site A, P26) |
| Facilitation (proposed elaboration) | Facilitation | Variable and under-resourced facilitation | Authority-sensitive facilitation | Facilitation in this setting was boundary-spanning work among intensive care, neurology, neurosurgery, nursing, and institutional leadership. It required someone credible across these groups, able to convene disagreement, negotiate ownership, and keep implementation from becoming a contest between services. A facilitator without standing could organize logistics but could not resolve the cross-professional disagreements on which adoption and sustainment depended. | a facilitator new to the program, "not having that what we call emotional currency" with leaders in the other services, "relied heavily on those of us who do have the emotional currency with those other leaders" (Site A, P21) |
| Facilitation; Context | Facilitation; Local level: Evaluation and feedback processes | Variable and under-resourced facilitation | Sustainment requires ongoing effort | Sustaining a change once introduced required active, ongoing effort rather than a single act of implementation, described as continuous audit and re-education. | "in God we trust[;] everyone else must bring data" (Site A, P31) |

i-PARIHS, integrated Promoting Action on Research Implementation in Health Services; PNCC, pediatric neurocritical care; PICU, pediatric intensive care unit.
